# Real-world Effect of Monoclonal Antibody Treatment in COVID-19 Patients in a Diverse Population in the United States

**DOI:** 10.1101/2021.04.08.21254705

**Authors:** Kaitlin Rainwater-Lovett, John T. Redd, Miles A. Stewart, Natalia Elías Calles, Tyler Cluff, Mike Fang, Mark J. Panaggio, Anastasia S. Lambrou, Jonathan K. Thornhill, Christopher Bradburne, Samuel Imbriale, Jeffrey D. Freeman, Michael Anderson, Robert Kadlec

**Author notes:** **Corresponding Author:** Kaitlin Rainwater-Lovett, PhD, MPH. **Alternate Corresponding Author:** John T. Redd, MD, MPH.

## Abstract

**Background:** Monoclonal antibodies (mAbs) against SARS-CoV-2 are a promising treatment for limiting the progression of COVID-19 and decreasing strain on hospitals. Their use, however, remains limited, particularly in disadvantaged populations.

**Methods:** Electronic health records were reviewed from SARS-CoV-2 patients at a single medical center in the United States that initiated mAb infusions in January 2021 with the support of the U.S. Department of Health and Human Services’ National Disaster Medical System. Patients who received mAbs were compared to untreated patients from the time period before mAb availability who met eligibility criteria for mAb treatment. We used logistic regression to measure the effect of mAb treatment on the risk of hospitalization or emergency department (E.D.) visit within 30 days of laboratory-confirmed COVID-19.

**Results:** Of 598 COVID-19 patients, 270 (45%) received bamlanivimab and 328 (55%) were untreated. Two hundred and thirty-one patients (39%) were Hispanic. Among treated patients, 5/270 (1.9%) presented to the E.D. or required hospitalization within 30 days of a positive SARS-CoV-2 test, compared to 39/328 (12%) untreated patients (p<0.001). After adjusting for age, gender, and comorbidities, the risk of E.D. visit or hospitalization was 82% lower in mAb-treated patients compared to untreated patients (95% confidence interval [CI]: 66%-94%).

**Conclusions:** In this diverse, real-world COVID-19 patient population, mAb treatment significantly decreased the risk of subsequent E.D. visit or hospitalization. Broader treatment with mAbs, including in disadvantaged patient populations, can decrease the burden on hospitals and should be facilitated in all populations in the United States to ensure health equity.

**Summary:** In a diverse, real-world COVID-19 patient population, treatment with monoclonal antibodies significantly decreased the risk of subsequent emergency department visit or hospitalization within 30 days of a positive SARS-CoV-2 viral test.

## BACKGROUND

In late 2019, a new respiratory infection was detected in China and alarmed global health experts with its growing case incidence and clinical severity [1,2]. Over the course of a few months, severe acute respiratory syndrome-coronavirus-2 (SARS-CoV-2) spread around the world, overwhelming health systems. While a substantial proportion of patients remain asymptomatic [3], coronavirus disease 2019 (COVID-19) can rapidly progress and require hospitalization and intensive care. Severe disease is associated with older age, obesity, and several chronic medical conditions including cardiovascular, kidney, and pulmonary comorbidities [4–7].

As of late January 2021, approximately 15,000 new COVID-19 hospital admissions were occurring per day in the United States (U.S.) and hospital bed capacity exceeded 72% [8,9]. As healthcare systems continued to approach maximum bed capacity, a critical need for therapeutic interventions to reduce COVID-related hospitalizations emerged. Although therapeutic options for COVID-19 remain limited, monoclonal antibodies (mAbs) that neutralize SARS-CoV-2 are a promising treatment for limiting the progression of disease. Four mAbs are available in the U.S. through Emergency Use Authorizations (EUAs) by the U.S. Food and Drug Administration (FDA): bamlanivimab monotherapy [10], bamlanivimab in combination with etesevimab [11], and casirivimab in combination with imdevimab [12]. These products are human IgG1 antibodies that neutralize the virus by binding the spike protein of SARS-CoV-2, preventing attachment of the virus to the human cellular receptor angiotensin-converting enzyme-2. A single infusion of bamlanivimab was recently demonstrated to reduce the risk of hospitalization, emergency department (E.D.) visits, and death among patients with mild to moderate COVID symptoms in randomized, controlled phase 2/3 trials by more than 70% [10].

Monoclonal antibodies are underutilized as a treatment for reducing severe disease and could significantly decrease hospitalizations and potentially long-term COVID effects [13]. Utilization can be particularly challenging in racial and ethnic minorities and disadvantaged populations, in whom prevalence of risk factors for COVID-19 progression and death may be higher. A recent review highlights racial and ethnic minorities are commonly employed in jobs that require in-person presence that increase exposure to SARS-CoV-2, language barriers that limit understanding of public health information, and poorer access to health care facilities [14]. These factors can delay treatment until patients are in a critical state, which can shorten the therapeutic window for effective mAb receipt or possibly preclude mAb use entirely. Thus, mAb may be particularly underutilized in precisely the populations that would have the greatest benefit, threatening to exacerbate existing health inequities in the United States. Two primary barriers to implementation of mAb infusion therapy at healthcare facilities are: 1) a limited understanding of the necessary resources and processes to mobilize infusion sites, and 2) understanding the magnitude of the potential impact of mAb treatment on reducing the severity of disease. We previously addressed the first barrier through a process assessment and improvement analysis [15], demonstrating considerable flexibility in assembling an infusion site and the feasibility of mAb delivery in diverse treatment locations. Here, we aim to determine the extent to which mAb treatment decreases COVID-related hospital admission and E.D. visits among patients with mild to moderate COVID-19 within 30 days of treatment in the U.S.

## METHODS

We conducted a retrospective cohort study in February 2021 of SARS-CoV-2-positive patients to evaluate the effect of mAb treatment on the risk of a medical visit within 30 days. This study evaluated patients who presented to a single medical center to which the U.S. Department of Health and Human Services’ Assistant Secretary for Preparedness and Response (ASPR) had deployed elements of the National Disaster Medical System (NDMS) to establish a mAb infusion site. This medical center is located in a moderately sized city with a population of approximately 500,000. The city’s population is 56.4% non-White with a median household income that is 64% of the U.S. level and a poverty rate of 23.4% [16,17]. This clinical support activity was conducted as part of the ASPR public health response to the COVID-19 pandemic and at the request of the host medical center. Under HHS Office of Health Research Protection guidelines, it was judged a non-research COVID-19 response [18]. The Johns Hopkins University Applied Physics Laboratory and the medical center concurred with a non-research determination.

The target population for this evaluation was patients with positive results of SARS-CoV-2 viral testing who were 12 years of age or older, at least 40 kg in weight, and at high risk for progressing to severe COVID-19 or hospitalization. Clinical data were obtained in February 2021 from electronic health records maintained by the medical system, which includes both a major medical center and several outpatient clinics with integrated health records.

Our retrospective cohort consisted of patients presenting to either outpatient clinics or the medical center who tested positive for SARS-CoV-2 via an antigen or polymerase chain reaction-based test. Patients with positive viral test results recorded in the electronic health record between July 1^st^ and December 20^th^, 2020 were identified as untreated patients. These patients were eligible for inclusion in the analysis if they met the eligibility criteria for mAb treatment (Table 1). Treatment with mAb became available at the medical center on January 7th, 2021. SARS-CoV-2 patients who received mAb infusions between January 7^th^ and January 15^th^, 2021, are referred to as treated patients. We selected the test date of December 20^th^, 2020, as the final date of eligibility for untreated patients to ensure no overlap in the treated and untreated patient populations based on the maximum ten-day symptom onset window permitting mAb treatment eligibility and decreased healthcare seeking behavior during winter holidays [19]. The decision to seek mAb treatment for COVID-19 was made by the patient and the provider. At presentation for mAb treatment, the date of SARS-CoV-2 test positivity was confirmed through paper records provided by the patient or rapid antigen test performed on-site, and intake staff collected demographic and clinical information, including eligibility criteria for treatment (Table 1). Any adverse events were recorded on patient forms. While mAb treatments continued after January 15^th^, the end date was established to permit sufficient follow-up at the time of data collection.

**Table 1.**
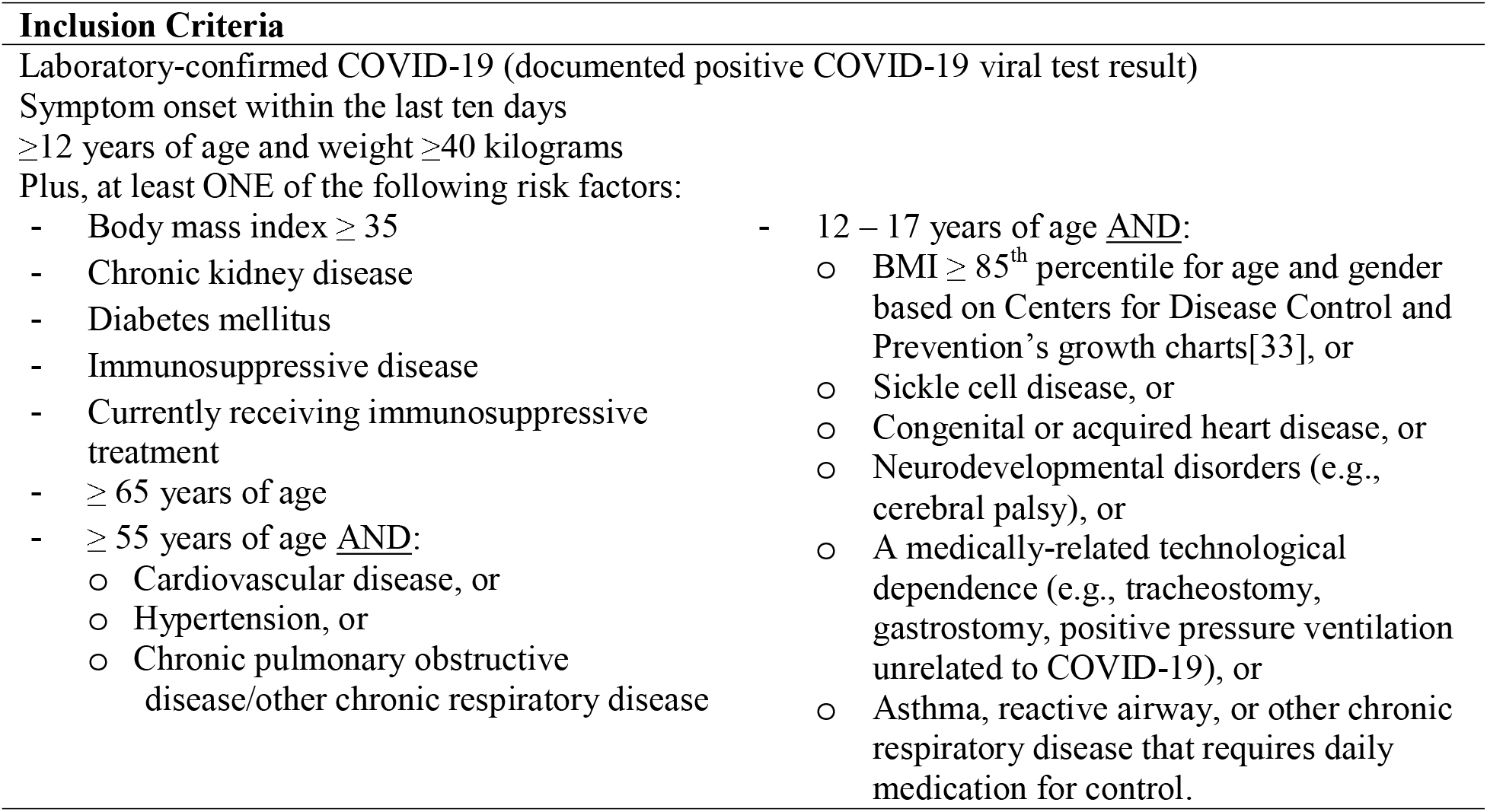
Eligibility criteria for SARS-CoV-2 monoclonal antibody infusions.

Data extracted from existing medical records included age, sex, race, ethnicity, height and weight, and presence of the following pre-existing conditions as recorded by clinicians in the health record: blood disorders (e.g., sickle cell disease, thalassemia), cancer, diabetes, Down syndrome, chronic lung disease, chronic liver disease, hypertension, immunosuppressive condition, chronic kidney disease, obesity or overweight, and organ transplant. Pre-existing conditions were captured from the Chief Complaint of health records within the six months prior to the date of SARS-CoV-2 testing. Laboratory values and clinical exam measurements were not extracted to define pre-existing conditions.

Race categories were defined as American Indian/Alaskan Native, Asian, Black, Hawaiian/Pacific Islander, White, and other. Ethnicity was defined as Hispanic or Non-Hispanic. Body mass index (BMI) was calculated as kilograms per meter-squared. In the absence of height and weight, the pre-existing conditions of “obesity” and “overweight” were used for BMI categorization. The composite outcome of a medical visit was defined as the first instance of COVID-19-related E.D. visit or hospitalization after positive SARS-CoV-2 viral test result and was obtained from the electronic health record. A medical visit was COVID-related if one or more of the following chief complaints were identified: blood in sputum, chest congestion, chest pain, cough, COVID-19 screening, difficulty breathing, fever, flu-like symptoms, hypoxia, shortness of breath, sore throat, or weakness [20–23]. Dates of COVID-19 symptom onset and positive SARS-CoV-2 antigen test results performed at the infusion center were recorded on paper-based forms upon arrival of patients for mAb treatment, but were not recorded in electronic health records.

Characteristics of patients were compared using Welch t-tests for continuous variables and Chi-squared test for categorical variables. Age was categorized as younger than or equal to 65 years of age or older than 65 years. Logistic regression was used to evaluate the effect of mAb treatment on medical visits that occurred within 30 days of SARS-CoV-2-positive viral test by applying a generalized linear model with a logits link function. The occurrence of a medical visit was evaluated as a binary outcome. Variables included in the model were those deemed epidemiologically relevant. Model diagnostics indicated that no data points substantially influenced model estimates, as assessed by Cook’s distance. All data processing and analyses were conducted using R version 4.0.3 [24].

## RESULTS

Medical records were available from 875 SARS-CoV-2-positive patients (Table 2) confirmed during July 1 through December 20, 2020. Of these, 547 patients did not meet eligibility criteria for mAb treatment (Table 1). This resulted in the analysis of 598 patients, 270 of whom (45%) were eligible for and received bamlanivimab during a single week in January 2021, comprising the treated group. A total of 328 untreated patients (55%) served as the historical comparator population. These untreated patients represented individuals who would have been eligible for mAb infusion had the treatment been available at the time of their COVID-19-positive viral test results.

**Table 2.** Baseline demographic and medical characteristics of SARS-CoV-2-positive patients. Abbreviations: Max, maximum; Min, minimum; SD, standard deviation.

|  | Untreated<br>(N=328) | mAB Treated<br>(N=270) | Overall<br>(N=598) | P-value |
| --- | --- | --- | --- | --- |
| **Sex** |  |  |  | 0.10 |
| Female | 211 (64.3%) | 155 (57.4%) | 366 (61.2%) |  |
| Male | 117 (35.7%) | 115 (42.6%) | 232 (38.8%) |  |
| **Age** |  |  |  | 0.03 |
| Mean (SD) | 61.0 (17.8) | 63.9 (15.9) | 62.3 (17.0) |  |
| Median [Min, Max] | 65.0 [13.0, 98.0] | 66.0 [18.0, 98.0] | 65.0 [13.0, 98.0] |  |
| **Age greater than 65** |  |  |  | 0.33 |
| Yes | 168 (51.2%) | 150 (55.6%) | 318 (53.2%) |  |
| No | 160 (48.8%) | 120 (44.4%) | 280 (46.8%) |  |
| **Race** |  |  |  | 0.66 |
| American Indian, Alaskan Native | 5 (1.5%) | 6 (2.2%) | 11 (1.8%) |  |
| Asian | 5 (1.5%) | 4 (1.5%) | 9 (1.5%) |  |
| Black | 14 (4.3%) | 5 (1.9%) | 19 (3.2%) |  |
| Hawaiian, Pacific Islander | 1 (0.3%) | 1 (0.4%) | 2 (0.3%) |  |
| Other | 11 (3.4%) | 6 (2.2%) | 17 (2.8%) |  |
| White | 278 (84.8%) | 214 (79.3%) | 492 (82.3%) |  |
| Missing | 14 (4.3%) | 34 (12.6%) | 48 (8.0%) |  |
| **Ethnicity** |  |  |  | 0.58 |
| Hispanic | 129 (39.3%) | 102 (37.8%) | 231 (38.6%) |  |
| Non-Hispanic | 188 (57.3%) | 133 (49.3%) | 321 (53.7%) |  |
| Missing | 11 (3.4%) | 35 (13.0%) | 46 (7.7%) |  |
| **BMI** |  |  |  | <0.01 |
| 30 or greater | 52 (15.9%) | 24 (8.9%) | 76 (12.7%) |  |
| 35 or greater | 35 (10.7%) | 11 (4.1%) | 46 (7.7%) |  |
| **Hypertension** |  |  |  | <0.01 |
| Yes | 176 (53.7%) | 55 (20.4%) | 231 (38.6%) |  |
| No | 152 (46.3%) | 215 (79.6%) | 367 (61.4%) |  |
| **Chronic Kidney Disease** |  |  |  | 0.22 |
| Yes | 19 (5.8%) | 9 (3.3%) | 28 (4.7%) |  |
| No | 309 (94.2%) | 261 (96.7%) | 570 (95.3%) |  |
| **Cardiovascular Disease** |  |  |  | <0.01 |
| Yes | 71 (21.6%) | 20 (7.4%) | 91 (15.2%) |  |
| No | 257 (78.4%) | 250 (92.6%) | 507 (84.8%) |  |
| **COVID-related ED visit or Admission within 30 days** |  |  |  | <0.01 |
| Yes | 39 (11.9%) | 5 (1.9%) | 44 (7.4%) |  |
| No | 289 (88.1%) | 265 (98.1%) | 554 (92.6%) |  |

Among the 598 patients, no statistically significant differences in sex or ethnicity were identified between the treated and untreated study groups (Table 2). Untreated patients were an average of three years younger than the treated patients (p=0.02), and health records were more likely to report untreated patients as overweight or obese and with a history of hypertension or cardiovascular disease (all p<0.001).

In the 30 days following a positive SARS-CoV-2 test result, five of 270 treated patients (1.9%) presented to the E.D. or required hospitalization within 30 days of a positive SARS-CoV-2 test result, compared to 39 of the 328 untreated patients (12%) (p<0.01) (Table 2). Untreated patients had a medical visit a median of four days after SARS-CoV-2-positive viral test result (interquartile range [IQR]: 2, 8 days), while treated patients had a medical visit an average of eight days after mAb treatment (IQR: 4, 8) (p=0.112 by Kolmogorov-Smirnov test). No adverse events were reported among mAb-infused patients.

Treatment with mAb was associated with an 82% decrease in the risk of a COVID-19-related medical visit within 30 days of a positive SARS-CoV-2 viral test after adjusting for demographic factors and pre-existing conditions (95% CI: 66%, 94%) (Table 3). A BMI ≥ 35 greatly increased the risk of a medical visit in the multivariable analysis (odds ratio: 6.44 [95% CI: 2.48, 16.71]). Age ≥ 65 was also associated with a 2.10-fold increased risk but this was not statistically significant (95% CI: 0.97, 4.77).

**Table 3.** Risk of COVID-19-related hospitalization or emergency department visit within 30 days of SARS-CoV-2-positive viral test. *p<0.05. Abbreviations: CI, confidence interval; mAb, monoclonal antibody; OR, odds ratio.

|  | **Unadjusted** |  | **Adjusted** |  |
| --- | --- | --- | --- | --- |
|  | OR | [95% CI] | OR | [95% CI] |
| (Intercept) |  |  | 0.05 | [0.02, 0.12] |
| **Gender** |  |  |  |  |
| Female | 1.00 | [0.54, 1.93] | 0.88 | [0.44, 1.78] |
| Male | *Reference* |  | *Reference* |  |
| **Age (years)** |  |  |  |  |
| ≤64 | *Reference* |  | *Reference* |  |
| >65 | 1.06 | [0.57, 1.99] | 2.10 | [0.97, 4.77] |
| **Race** |  |  |  |  |
| Black | 1.60 | [0.23, 5.90] | 1.2 | [0.17, 5.12] |
| White | *Reference* |  | *Reference* |  |
| Other | 0.36 | [0.02, 1.71] | 0.35 | [0.02, 1.79] |
| Unknown | 0.29 | [0.01, 1.35] | 0.54 | [0.03, 2.87] |
| **Ethnicity** |  |  |  |  |
| Hispanic | 1.50 | [0.80, 2.78] | 1.66 | [0.84, 3.32] |
| Non-Hispanic | *Reference* |  | *Reference* |  |
| **Body Mass Index** |  |  |  |  |
| <30 | *Reference* |  | *Reference* |  |
| ≥30 and <35 | 1.61 | [0.67, 3.46] | 1.98 | [0.76, 4.76] |
| ≥35 | 4.95 | [2.21, 10.5]* | 6.44 | [2.48, 16.71]* |
| **Comorbidities** |  |  |  |  |
| Hypertension | 2.22 | [1.19, 4.19]* | 1.37 | [0.67, 2.81] |
| Chronic Kidney Disease | 1.61 | [0.36, 4.89] | 1.15 | [0.25, 3.79] |
| Cardiovascular Disease | 1.73 | [0.78, 3.54] | 1.07 | [0.45, 2.40] |
| **mAb Treatment** | 0.14 | [0.05, 0.34]* | 0.18 | [0.06, 0.44]* |

## DISCUSSION

This study demonstrated that a single infusion of bamlanivimab within 10 days of COVID-19 symptom onset decreased the risk of COVID-related hospitalization and E.D. visits among a real-world, diverse patient population in the U.S. who were at risk of progression to severe disease compared with an historical untreated population. The association between treatment and improved clinical outcome remained significant after controlling for gender, age, race, ethnicity, and pre-existing conditions. A BMI of greater than 35 remained highly associated with disease progression requiring a medical visit after adjusting for mAb treatment and other co-factors.

Approximately 2% of the treated group were hospitalized or visited the E.D. after mAb infusion, which was similar to the rate of medical visits in the efficacy assessment of bamlanivimab [10]. In contrast, almost 12% of untreated patients in the current study required a medical visit within 30 days of a positive COVID test. This risk was nearly double the 6.3% of placebo controls who presented to the E.D., required hospitalization, or died in the Phase 2/3 trial [10], suggesting that the current study’s patient population was older and had a higher risk of progression to severe disease. This difference reinforces the need to evaluate therapeutics in diverse populations and in real-world clinical situations, as patients who are referred for and receive treatments often differ from those who are enrolled in a clinical trial.

Few treatment options have been available during the COVID-19 pandemic for reducing the severity of disease and preventing hospitalization, leading to significant strain on many hospitals[8]. Reducing the proportion of patients who progress to severe disease and require hospitalization by approximately 80% would be of immense value to medical centers, in which intensive care units contain an average of only 15 staffed beds [25]. The availability of mAbs at no drug cost due to their procurement by the U.S. government places a therapeutic option more easily within reach of many who are at the highest risk of severe disease.

The use of electronic health records is a strength of the current study. Due to the medical center’s electronic record system, we were able to assemble a SARS-CoV-2-positive cohort who would have been eligible for mAb treatment at the time of their diagnosis based on pre-existing risk factors, had the therapeutics been available at that time. An additional strength of this study was the diverse patient population in the area, resulting in the inclusion of a large proportion of patients of Hispanic ethnicity (39%). Our results are consistent with prior clinical trial data showing a 70% reduction in medical visits by mAb-infused patients compared to placebo controls[10]. A BMI of 35 or higher was a strong independent predictor of an increased risk of medical visits, which was consistent with other COVID-19 studies [7].

A significantly larger proportion of untreated patients had co-morbidities that increase the risk of severe COVID-19 outcomes compared to treated patients in the current study, notably a higher proportion with elevated BMI. Although mAb treatment remained significantly associated with a decreased risk of hospitalization or E.D. visit after adjusting for pre-existing conditions (82% reduction; 95% CI: 66%, 94%), the baseline differences between the treated and untreated groups suggest a potential difference in accessibility of mAb treatment. For example, patients with fewer co-morbidities may have more easily been able to avail themselves of treatment. The continued U.S. government efforts to increase access to mAbs are intended to ensure that COVID-19 therapeutics are equally available to all patients - an important national health equity consideration.

To receive mAb infusions, patients must seek out treatment within 10 days of a positive SARS-CoV-2 antigen test result. This can be burdensome and stresses the importance of widespread availability of testing. Evidence also suggests that patients with more significant or severe co-morbidities are likely to have more complete health records [26,27]. This effect may have overrepresented patients with more severe chronic conditions into the untreated group based on the application of mAb eligibility criteria for inclusion in the analysis. Additionally, without active follow-up of patient outcomes, misclassification of the medical visit outcome was possible as patients could seek follow-up care at any facility. These considerations and the differences between the study groups suggest confounders remain that were unmeasured in this analysis and may reflect the retrospective untreated population group in the study’s design. These limitations could be further evaluated in a larger, prospective, observational study.

While individuals at increased risk of SARS-CoV-2 infection and severe disease are prioritized for vaccination in most U.S. states, therapeutic options such as mAb infusions remain a necessity for those who remain unvaccinated due to contraindications or vaccine hesitancy [28,29]. Although viral variants are being discovered that are poorly neutralized by several mAbs in laboratory studies [30,31], suggesting reduced effectiveness in patient populations, relatively minor adjustments to the currently available mAb products can counter these changes. Additionally, the FDA has issued guidance encouraging use of existing formulations, platforms, and clinical protocols to facilitate expedited review and rapid introduction of these modified mAb products to general public [32].

In summary, we demonstrated that mAb treatment with bamlanivimab was associated with an approximately 80% reduction in the risk of medical visits among a diverse COVID-19 patient population under real-world conditions. Increasing availability and utilization of novel COVID-19 therapeutics may improve patient outcomes, reduce burden on the health system, and contribute to increased health equity in the United States.

## Data Availability

The data was used from electronic medical records as part of process improvement and is not publicly available.

## FUNDING AND CONFLICT OF INTEREST STATEMENT

All authors have completed the ICMJE uniform disclosure form: all authors had financial support from the U.S. Department of Health and Human Services, Office of the Assistant Secretary for Preparedness and Response for the submitted work; no financial relationships with any organizations that might have an interest in the submitted work in the previous three years; no other relationships or activities that could appear to have influenced the submitted work.

## ACKNOWLEDGEMENTS

The authors acknowledge the significant efforts of George (Mark) Thorp, RN, EMT-P, who led and coordinated the Disaster Medical Assistance Team’s mAb infusion site set-up, initiation, and integration with the Tucson Medical Center. The authors thank David W. Forest, Christopher Florko, and Judy McCord from TMC Health Care, who provided extensive time and support for this analysis.

## FIGURES AND TABLES

**Figure 1.**
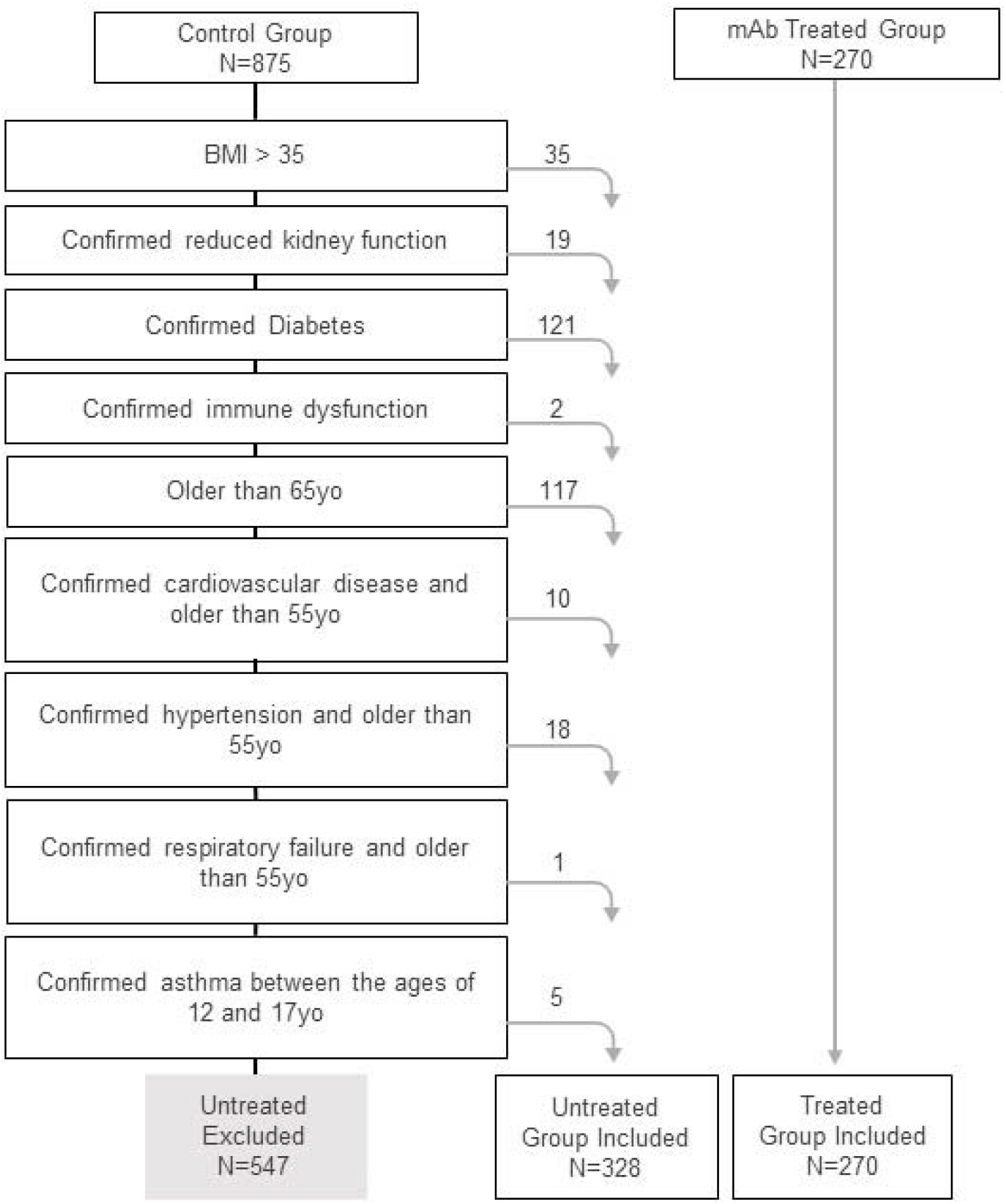
Flow diagram applying the inclusion criteria to collected health records that generated the final study population.

**Supplemental Table.** STROBE Statement

|  | **Item No** | **Recommendation** | **Page No** |
| --- | --- | --- | --- |
| **Title and abstract** | 1 | (a) Indicate the study's design with a commonly used term in the title or the abstract,<br>(b) Provide in the abstract an informative and balanced summary of what was done and what was found | Pg 3 |
| **Introduction** |  |  |  |
| Background/rationale | 2 | Explain the scientific background and rationale for the investigation being reported | Pgs 4-5 |
| Objectives | 3 | State specific objectives, including any prespecified hypotheses | Pg 5 |
| **Methods** |  |  |  |
| Study design | 4 | Present key elements of study design early in the paper | Pgs 5-6 |
| Setting | 5 | Describe the setting, locations, and relevant dates, including periods of recruitment, exposure, follow-up, and data collection | Pgs 5-6 |
| Participants | 6 | (a) Give the eligibility criteria, and the sources and methods of selection of participants. Describe methods of follow-up<br>(b) For matched studies, give matching criteria and number of exposed and unexposed | Pg 6 |
| Variables | 7 | Clearly define all outcomes, exposures, predictors, potential confounders, and effect modifiers. Give diagnostic criteria, if applicable | Pgs 6-7 |
| Data sources/<br>measurement | 8* | For each variable of interest, give sources of data and details of methods of assessment (measurement). Describe comparability of assessment methods if there is more than one group | Pgs 6-7 |
| Bias | 9 | Describe any efforts to address potential sources of bias | Pg 6 |
| Study size | 10 | Explain how the study size was arrived at | Pgs 6-7 |
| Quantitative variables | 11 | Explain how quantitative variables were handled in the analyses. If applicable, describe which groupings were chosen and why | Pgs 7-8 |
| Statistical methods | 12 | (a) Describe all statistical methods, including those used to control for confounding<br>(b) Describe any methods used to examine subgroups and interactions<br>(c) Explain how missing data were addressed<br>(d) If applicable, explain how loss to follow-up was addressed<br>(e) Describe any sensitivity analyses | Pg 8 |
| **Results** |  |  |  |
| Participants | 13* | (a) Report numbers of individuals at each stage of study—eg numbers potentially eligible, examined for eligibility, confirmed eligible, included in the study, completing follow-up, and analysed<br>(b) Give reasons for non-participation at each stage<br>(c) Consider use of a flow diagram | Pg 8 |
| Descriptive data | 14* | (a) Give characteristics of study participants (eg demographic, clinical, social) and information on exposures and potential confounders<br>(b) Indicate number of participants with missing data for each variable of interest<br>(c) Summarise follow-up time (eg, average and total amount) | Pg 8 |
| Outcome data | 15* | Report numbers of outcome events or summary measures over time | Pg 9 |
| Main results | 16 | (a) Give unadjusted estimates and, if applicable, confounder-adjusted estimates and their precision (eg, 95% confidence interval). Make clear which confounders were adjusted for and why they were included<br>(b) Report category boundaries when continuous variables were categorized<br>(c) If relevant, consider translating estimates of relative risk into absolute risk for a meaningful time period | Pg 9 |
| Other analyses | 17 | Report other analyses done—eg analyses of subgroups and interactions, and sensitivity analyses | N/A |
| **Discussion** |  |  |  |
| Key results | 18 | Summarise key results with reference to study objectives | Pgs 9-12 |
| Limitations | 19 | Discuss limitations of the study, taking into account sources of potential bias or imprecision. Discuss both direction and magnitude of any potential bias | Pg 11 |
| Interpretation | 20 | Give a cautious overall interpretation of results considering objectives, limitations, multiplicity of analyses, results from similar studies, and other relevant evidence | Pg 12 |
| Generalisability | 21 | Discuss the generalisability (external validity) of the study results | Pgs 9, 12 |
| **Other information** |  |  |  |
| Funding | 22 | Give the source of funding and the role of the funders for the present study and, if applicable, for the original study on which the present article is based | Pg 12 |

## Notes

### Competing Interest Statement

The authors have declared no competing interest.

### Author Declarations

This clinical support activity was conducted as part of the Office of the Assistant Secretary of Preparedness and Response's public health response to the COVID-19 pandemic and at the request of the host medical center. Under the Department of Health and Human Services Office of Health Research Protection guidelines, it was judged a non-research COVID-19 response. The Johns Hopkins University Applied Physics Laboratory and the medical center concurred with a non-research determination.

